# Estimating the Effect of Social Distancing Interventions on COVID-19 in the United States

**DOI:** 10.1101/2020.07.10.20151001

**Authors:** Andrew M. Olney, Jesse Smith, Saunak Sen, Fridtjof Thomas, H. Juliette T. Unwin

## Abstract

Since its global emergence in 2020, severe acute respiratory syndrome coronavirus 2 (SARS-CoV-2) has caused multiple epidemics in the United States. Because medical treatments for the virus are still emerging and a vaccine is not yet available, state and local governments have sought to limit its spread by enacting various social distancing interventions such as school closures and lockdown, but the effectiveness of these interventions is unknown. We applied an established, semi-mechanistic Bayesian hierarchical model of these interventions on SARS-CoV-2 spread in Europe to the United States. We estimated the effect of interventions across all states, contrasted the estimated reproduction number, R_t_, for each state before and after lockdown, and contrasted predicted future fatalities with actual fatalities as a check on the model’s validity. Overall, school closures and lockdown are the only interventions modeled that have a reliable impact on R_t_, and lockdown appears to have played a key role in reducing R_t_ below 1.0. We conclude that reversal of lockdown, without implementation of additional, equally effective interventions, will enable continued, sustained transmission of SARS-CoV-2 in the United States.

## 1. Introduction

Severe acute respiratory syndrome coronavirus 2 (SARS-CoV-2) causes coronavirus disease 2019 (COVID-19). Initially discovered in Wuhan, China in December 2019, SARS-CoV-2 rapidly spread to the rest of the world, initially through travelers from Wuhan, but later through community transmission in Asia, Europe, Australia, and North America, until it was declared a pandemic by the World Health Organization on March 11, 2020. The rapid spread of SARS-CoV-2 is attributable to its transmissibility by aerosol and fomites (van Doremalen et al., 2020; Bourouiba, 2020), by presymptomatic/asymptomatic carriers (Bai et al., 2020; Furukawa et al., 2020), and by the relatively mild clinical characteristics of symptomatic carriers, which often include fever, cough, and fatigue (Guan et al., 2020). However, approximately 20% of confirmed cases develop severe or critical forms of COVID-19, including complications of respiratory failure, myocardial dysfunction, and acute kidney injury, with approximiately 50% mortality for critically-ill patients (Phua et al., 2020).

As of July 2020, outbreaks or epidemics of SARS-CoV-2 have emerged in all 50 states, with over 2.5 million confirmed cases reported. Because medical treatments for and vaccinations against the virus are still emerging, state and local governments have sought to limit its spread by enacting various social distancing interventions. Social distancing interventions have varied widely both within states and across states. Within states, interventions typically have begun with public health directives like washing hands and staying home if sick, followed by restrictions on or closures of places housing vulnerable populations like nursing homes or schools, followed by successive, increasingly restrictive bans on gathering in groups, culminating in stay-at-home orders or so-called lockdown. Across states, interventions have been adopted with different speeds, such that some states moved rapidly to lockdown and other states never entered lockdown at all. Likewise, states are currently lifting lockdown and reversing social distancing interventions at different rates.

To explore the effect of social distancing interventions, we applied an established, semi-mechanistic Bayesian hierarchical model of these interventions on SARS-CoV-2 spread in Europe (Flaxman et al., 2020a; Flaxman et al., 2020b) to the United States. We estimated the effect of interventions and the time-varying reproduction number (R_t_) for each state using state-level daily case fatality counts.

## 2. Methods

### 2.1. Data

In this study, we used data from three different sources: state-level intervention data, infection fatality rate data, and confirmed case fatality data.

#### State-level intervention data

We created a dataset of state-level intervention dates by inspecting the executive orders, public health directives, and official communications (e.g., press releases) from state governments (Olney and Olney, 2020). For each intervention date, we used the effective date, unless the timing of the intervention was so close to midnight as to only practically have effect the next day. Interventions were only counted if they targeted the general population, e.g., restricting out-of-state travel by state employees only was not counted as a travel restriction intervention. All interventions dates are linked to their source documents with any needed commentary for data provenance. The interventions themselves closely parallel those in the European model we used, but with slightly different operationalizations which we describe in turn. *Self isolating if ill* is a recommendation to stay home if sick. *Social distancing encouraged* is a recommendation to avoid nonessential travel and/or contact; the mere words “social distancing” were not counted unless they were elaborated with examples of what social distancing entails. *Schools or universities closing* is the date at which schools partly or completely close; the earlier of schools or universities closing was used. *Sport* is the banning of sporting events or public gatherings of more than 1000 persons. *Public events* is the banning of public gatherings of more than 100 participants. Finally, *lockdown* includes banning of non-essential gatherings or business operations, which is sometimes formalized as a stay-at-home or safer-at-home order. Notably some more restrictive interventions imply others, e.g., *lockdown* implies all other interventions, and *public events* implies *sport*.

#### Infection fatality rate data

The infection fatality rate (IFR), or ratio of fatalities to true infections, was derived via the methods outlined in Flaxman et al. Briefly, IFR estimates from Verity et al. (2020) were adjusted using an age-specific UK contact matrix to account for non-uniform attack rates across age groups (see Ferguson et al. 2020 for details and previous US application). The resulting IFRs were weighted by state-level age demographics and averaged to produce estimates adjusted for both age and location. Demographic data were obtained from the 2018 ACS survey 5-year estimates (U.S. Census Bureau, 2020).

#### Confirmed case fatality data

SARS-CoV-2 fatality data was obtained from the New York Times public data repository (The New York Times, 2020), which describes the data collection process along with subtle issues in counting cases, e.g. cruise ship passengers. In general, the dataset counts confirmed cases according to where they were treated. Deaths are counted on the days they were reported up to midnight Eastern Time. Because this dataset provides cumulative counts, we transformed these into daily counts by taking the difference between successive daily cumulative counts (setting this difference to zero in the rare instances where cumulative counts were decreasing due to reporting corrections).

## 3. Model

We applied an established, semi-mechanistic Bayesian hierarchical model of interventions on SARS-CoV-2 spread in Europe to the United States, and the design and details of this model are presented elsewhere (Flaxman et al., 2020a; Flaxman et al., 2020b). Notably, a recent variant of this model has been applied to the United States at the state level, but this variant uses mobility data rather than interventions as the basis of predictions (Unwin et al., 2020). Briefly stated, daily death counts in the model follow a negative binomial distribution such that their expectation is a function of infections on previous days. The model is semi-mechanistic in the sense that it incorporates classical Susceptible-Infected-Removed concepts (Horsburgh and Mahon, 2008) in a Bayesian framework. The number of infected is modeled using a discrete renewal process that accounts for population saturation. Death counts are similarly linked to the number of infected based on the country (or state in the present case) IFR and the distribution of times from infection to death. Importantly, the model assumes the effect of intervention is that same regardless of location and that the implementation of an intervention instantaneously reduces R_t_. Making these assumptions allows data from multiple locations to be pooled for estimating intervention effects. The model was specified using Stan (Carpenter et al., 2017), and model inference was performed using adaptive Hamiltonian Monte Carlo. We fit our model with a time series for each state 30 days before the state has experienced seven deaths^1^, up to April 25, 2020, when some states began reversing their interventions.

## 4. Results

The mean number of days between the first and last intervention of a state was 19.53 days (SD = 5.73, range: 8-31). While each of 50 states had the opportunity to implement six different social distancing interventions, only 289 or 96.33% were implemented, with lockdown being the least implemented (n = 43). The mean IFR was 1.11% (SD = .12%, range: .76 - 1.35%). Because confirmed case fatality data increased dramatically over the time period examined, similar statistics are not reported for these data.

Estimated national intervention effects on R_t_ are shown in Table 1. Each estimated effect is an absolute reduction in R_t_. It is evident that only schools or universities closing and lockdown have a nontrivial impact on R_t_. Moreover, schools or universities closing and lockdown are the only interventions whose 95% credible interval does not cross zero.

**Table 1:** National intervention effects on R_t_.

| Intervention | Mean | 95% CI |  |
| --- | --- | --- | --- |
|  |  | LL | UL |
| Self isolating if ill | .012 | .000 | .059 |
| Sport | .021 | .000 | .102 |
| Social distancing encouraged | .033 | .000 | .162 |
| Public events | .103 | .000 | .379 |
| Schools or universities closing | .271 | .007 | .518 |
| Lockdown | .785 | .592 | .987 |

*Note.* Abbreviations: CI, credible interval; LL, lower limit; UL, upper limit.

State-level measures and estimates of the model are shown in Table 2. Of primary interest are the R_t_ estimates before and after lockdown and the corresponding death counts 2 weeks into the future, which are contrasted with actual deaths to assess model validity. Notably, no state had a mean R_t_ below 1.0 pre-lockdown, but 29 states had a R_t_ below 1.0 after lockdown. While lockdown had a strong effect in reducing R_t_ in all states that underwent lockdown, in these 29 states, lockdown appears to have been the single critical intervention allowing containment of the disease.

**Table 2.**
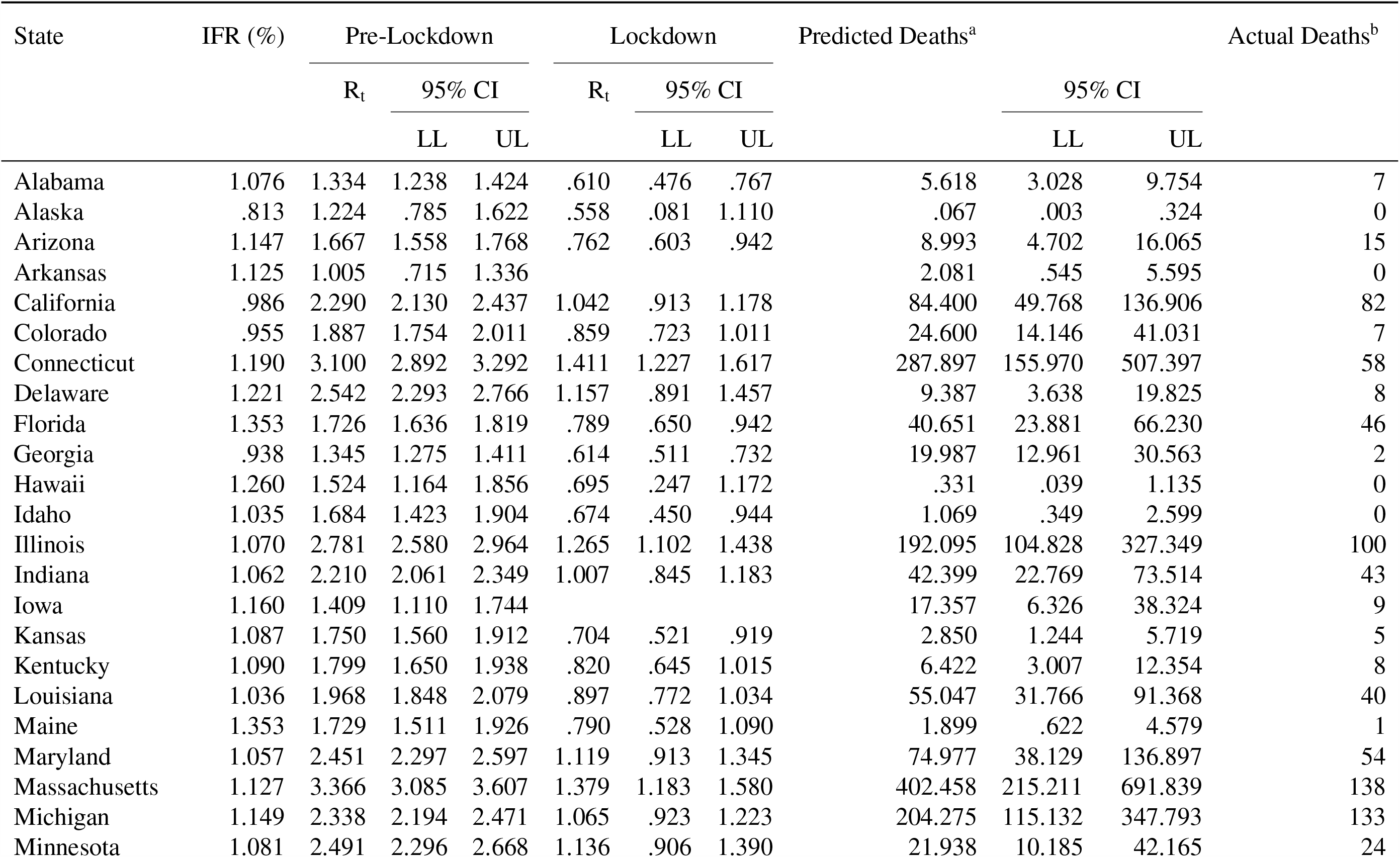
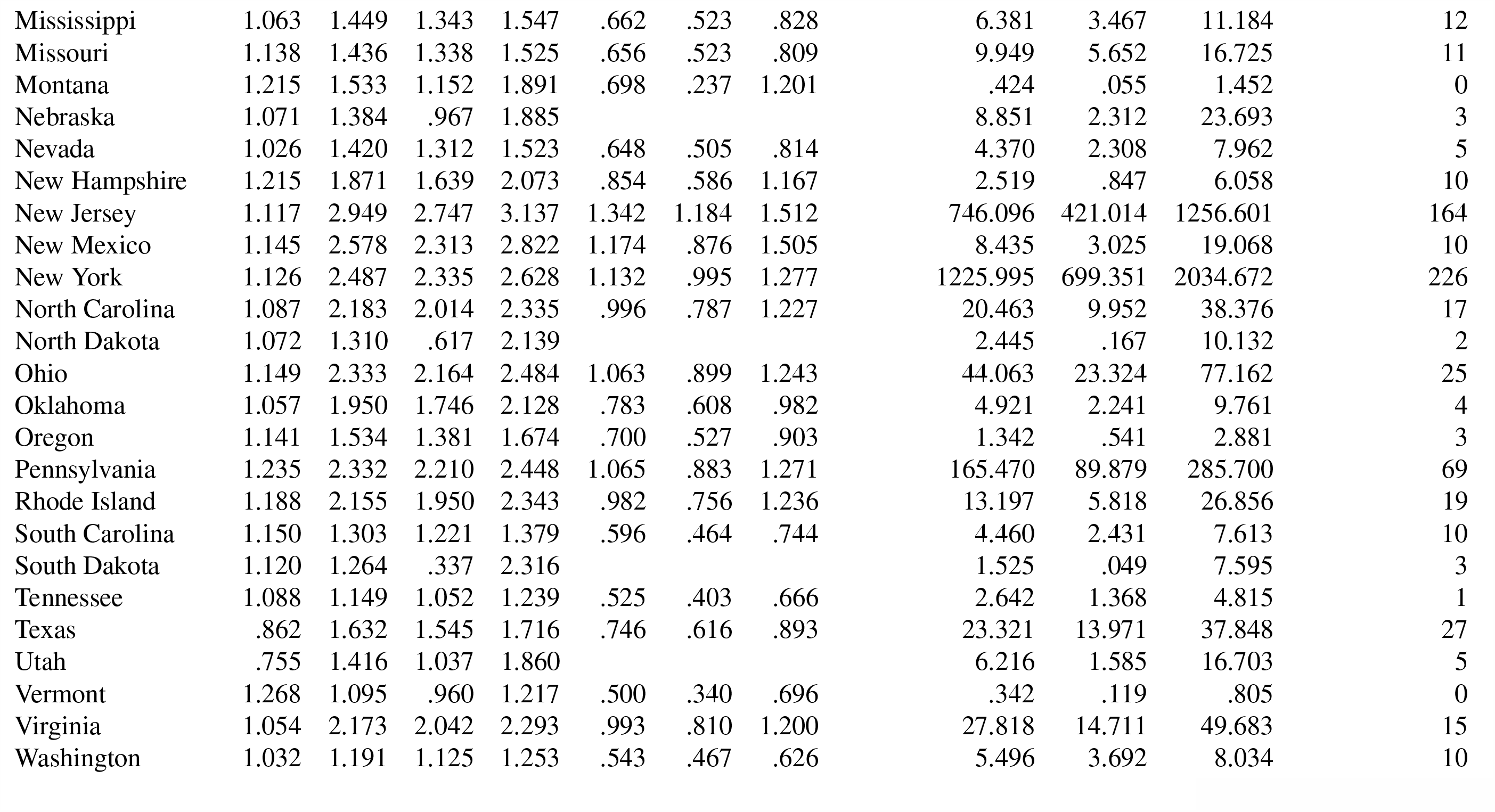
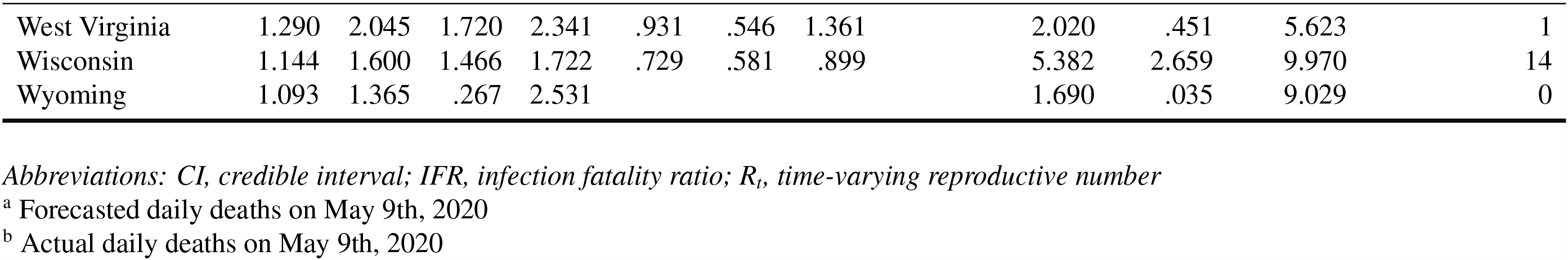
State-level measures and estimates of infection, fatality, and lockdown intervention.

Predicted deaths vs. actual deaths in each state serve as a validity check on the model’s estimates of intervention effects. Thirty-six states (72%) had actual deaths that were within the 95% CI of predicted deaths. Notably, the mean predicted deaths were well above actual (*>*100 deaths) for Connecticut, New Jersey, Massachusetts, and New York. The mean absolute error of mean predicted deaths was 50.80, and without these four states the mean absolute error was 10.08.

## 5. Discussion

Social distancing interventions are important for limiting the spread of SARS-CoV-2, because medical treatments for COVID-19 are still emerging and a vaccine is not available. To our knowledge, we are the first to apply an established, semi-mechanistic Bayesian hierarchical model of these interventions on SARS-CoV-2 spread in Europe to the United States. We estimated the effect of interventions across all states, contrasted the estimated R_t_ for each state before and after lockdown, and contrasted predicted future fatalities with actual fatalities as a check on the model’s validity. Overall, school closures and lockdown orders are the only interventions modeled that have an estimated effect where the 95% credible interval does not include zero, i.e. no effect. No state had an estimated R_t_ below 1.0 before lockdown, but 29 states reached an R_t_ below 1.0 after lockdown. The model’s ability to successfully predict future deaths supports the validity of estimated intervention effects. These results suggest that reversal of lockdown, without implementation of additional, equally effective interventions, will enable continued, sustained transmission of SARS-CoV-2 in the United States.

Our study has several limitations. First, the assumption that all interventions have the same implementation and effect in all states is a strong assumption. For example, states with a stronger culture of recreational sports will likely see a greater impact of the sports intervention than states without that culture. More directly, the intervention banning public gatherings of 100 persons or more could be met by a ban on 10 persons or more or 50 persons or more; it is unlikely that such bans are truly equivalent. This limitation has since been partially addressed in the European model by allowing random effects for lockdown only. Second, the assumption that interventions are binary rather than continuously varying is also a strong assumption and clearly an oversimplification, because it does not account for time-varying compliance with intervention. Several groups are incorporating mobility data as a measure of population mixing (Unwin et al., 2020; Woody et al., 2020; Team and Murray, 2020). Third, the parameters of the model are estimated using reasonable, but still uncertain, assumptions about prior distributions. We have used the same assumptions as in the European model, but these assumptions may be contradicted by future empirical work.

Modeling of SARS-CoV-2 is emerging and rapidly diversifying, including classical SEIR models and derivatives (Pei and Shaman, 2020), deep learning (Prakash, 2020), and piecewise models for sub-exponential growth (Scire et al., 2020). State and local governments are likewise rapidly adjusting their policy decisions regarding interventions based on case data and economic concerns. As the United States adopts an increasingly fragmented response to SARS-CoV-2, modeling approaches like ours that focus on shared interventions may not be tenable. While our results give valuable insights into which interventions did and which did not change the transmission rate substantially, we recommend that future studies measure the change in behaviors resulting from interventions and then strengthen the predictive relationships between these behaviors and disease transmission.

## Data Availability

Links to data are in the references

https://zenodo.org/record/3901617

## Footnotes

1 The Europe model used ten deaths as a somewhat arbitrary threshold for excluding imported cases; seven is the highest number we can use and still obtain valid data for states like Alaska, which have relatively low case count, cf. Unwin et al. (2020).

